# *N*-glycans mannosylation controls T-cell development by reprogramming thymocyte commitment, fate and repertoire

**DOI:** 10.1101/2022.02.04.22270265

**Authors:** Manuel M Vicente, Inês Alves, Ângela Fernandes, Ana M Dias, Elena Pérez-Anton, Alexandra Correia, Afonso R M Almeida, Gabriel A Rabinovich, Manuel Vilanova, Ana E Sousa, Salomé S Pinho

**Affiliations:** i3S – Institute for Research and Innovation in Health, University of Porto, 4200-135 Porto, Portugal; Institute of Biomedical Sciences Abel Salazar (ICBAS), University of Porto, 4050-313 Porto, Portugal; Graduate Program in Areas of Basic and Applied Biology (GABBA), ICBAS, University of Porto, 4050-313 Porto, Portugal; Faculty of Medicine, University of Porto, 4200-319 Porto, Portugal; Instituto de Medicina Molecular João Lobo Antunes, Faculdade de Medicina, Universidade de Lisboa, 1649-028 Lisboa, Portugal; Laboratorio de Inmunopatología, Instituto de Biología y Medicina Experimental (IBYME), Consejo Nacional de Investigaciones Científicas y Técnicas (CONICET), 1428 Ciudad de Buenos Aires, Argentina; Laboratorio de Inmuno-oncología Translacional, Instituto de Biología y Medicina Experimental (IBYME), Consejo Nacional de Investigaciones Científicas y Técnicas (CONICET), 1428 Ciudad de Buenos Aires, Argentina; Facultad de Ciencias Exactas y Naturales (FCEyN), Universidad de Buenos Aires, 1428 Ciudad de Buenos Aires, Argentina

**Author notes:** Corresponding author (S.S.P).

## Abstract

T-cell development is a major physiological process occurring in complex organisms, that ensures the formation of T-cell receptors diverse repertoire, to recognize antigens throughout the life of the organism. The thymus offers a microenvironment for efficient development, where progenitor populations go through maturation steps, which, when not regulated, are associated with disease onset, including autoimmune disorders and cancer. Glycosylation is a major post-translational modification that occurs in virtually all cells, including T lymphocytes, regulating receptor- turnover, affinity and signaling. However, there is a missing knowledge on how glycans regulate lymphocyte development and their impact in T-cell functions. We discovered stage-specific glycosylation profiles in human and murine thymocyte populations. Thereafter, we generated two glycoengineered mouse models displaying *N*-glycosylation pathway deficiencies, at early DN stages, lacking the *Mgat1* or *Mgat2* genes. We demonstrated remarkable defects in key T-cell developmental stages, such as ß- and DP-selection, natural regulatory T-cell generation, *γδ* T development/differentiation and thymic egression, in *Mgat1*-deficient thymocytes, indicating a pathogenic role of mannose *N*-glycans in development. We also demonstrated that a single *N*- glycan antenna (modelled in *Mgat2*-deficient thymocytes) is sufficient to rescue key developmental processes. In conclusion, we demonstrated that mannosylated thymocytes render a dysregulation in T-cell development, associated with disease.

**Graphical abstract:** 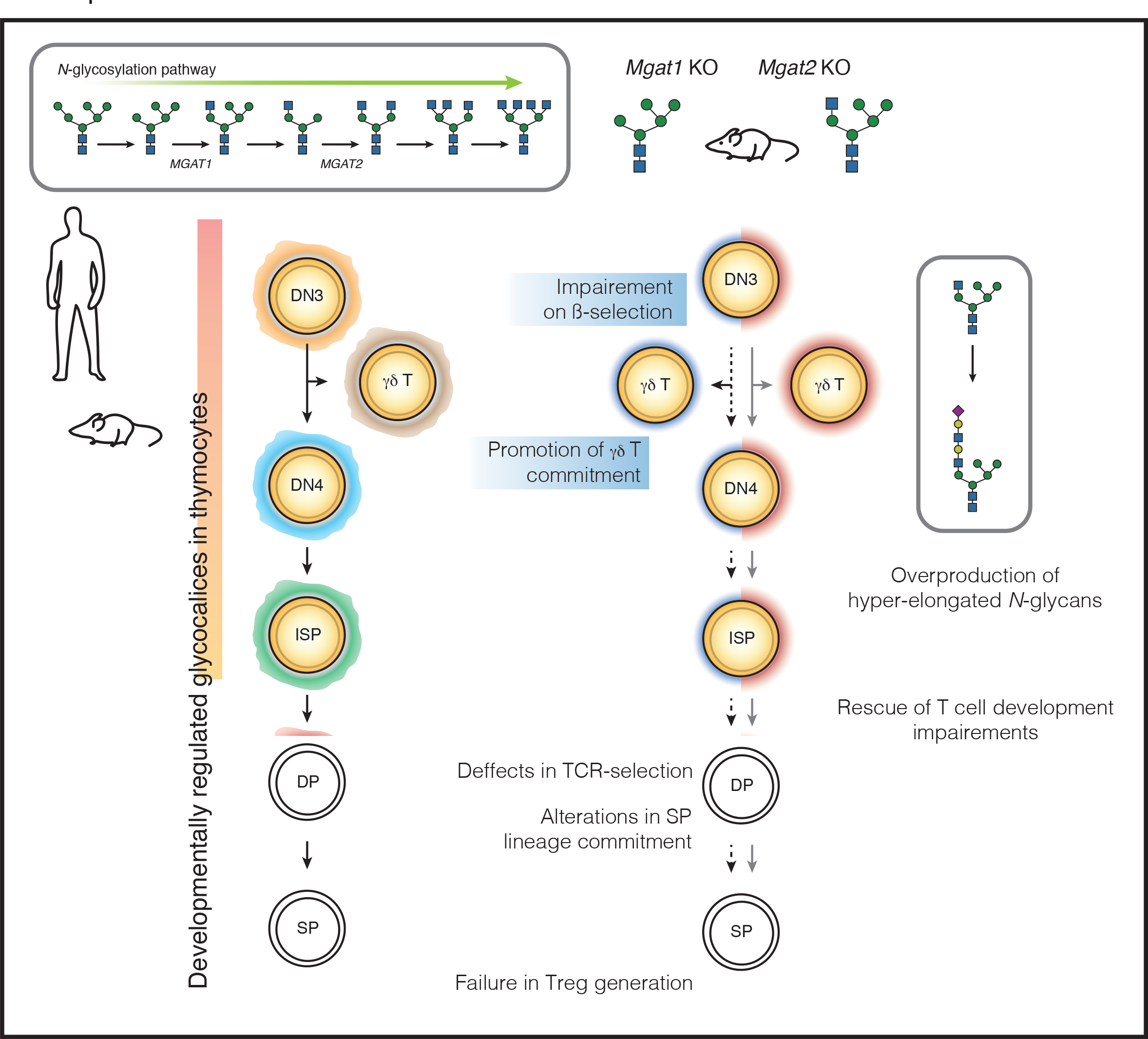

## INTRODUCTION

T-cell development is a tightly regulated process that occurs in the thymus, ensuring the formation of a T-cell pool with a diverse repertoire of T-cell receptors (TCRs), essential in adaptive immunity[1–3] Thymocytes enter through a series of developmental stages, mainly defined by variations in coreceptors expression. In the CD4^-^CD8^-^ double-negative-3 (DN3) stage, cells commit to the αß or the *γδ* lineages. Then massive proliferation of DN4 cells leads to the generation of CD4^+^CD8^+^ double-positive (DP) thymocytes, where a successful rearrangement of the *Tcra* locus generates a mature- and cell-unique TCR. An intermediate population is found on the DN4- to-DP transition, the CD8^+^CD3^-^ (mice) or CD4^+^CD3^-^ (humans) immature-single-positive (ISP) cells. Development of DP into mature CD4^+^ or CD8^+^ single-positive (SP) thymocytes is highly regulated by two selection steps: “positive-” and “negative-selection”, where non- or self-reactive T-cells are eliminated. Moreover, cells with TCR affinities just below the threshold for negative selection, develop into natural-regulatory T-cells (nTregs)[4, 5].

Glycans are present in essentially all cellular surfaces, being an important regulator of the immune system. In fact, T-cells contain a dense coat of glycans (glycocalyx) that tightly regulate cell activity and function[6–8]. Glycosylation is the enzymatic process responsible for the attachment of glycans to proteins/lipids by a portfolio of glycosyltransferases and glycosidases, acting in a step-wise manner[9]. Protein glycosylation has been shown to be essential in the regulation of T-cell maturation, activation and differentiation[10, 11]. However, its impact in T-cell development and in disease susceptibility remains poorly understood. Previous evidences have demonstrated essential contributions of *α*2,3-linked sialic-acid to T-cell development[12].

Furthermore, *N*-glycan branching was also shown to regulate DP selection[13]. In fact, complex-branched *N*-glycans have been described to be chief regulators of T-cell functions[6, 10].

Specifically, we and other have shown that the deficiency of β1,6-GlcNAc-branched complex *N*- glycans promote TCR clustering and signaling, resulting in lower activation thresholds, associated with increased susceptibility to multiple sclerosis and inflammatory bowel disease (IBD)[7,14-16].

Overall, these evidences support the importance of *N*-glycans as key regulators of peripheral T-cell activity and function, but the knowledge of their role in central tolerance and T-cell development is still unclear. Particularly, the glycome profiles of thymocytes along development and the impact of glycosylation changes in disease susceptibility remain largely unexplored. Here, we showed that both human and murine thymocyte populations display unique and distinct glycosylation signatures that define developmental stages. We further demonstrated that the premature truncation of the *N*-glycosylation pathway, early in murine T-cell development. globally impairs their development, resembling a primary immunodeficient-like syndrome, with lower number of peripheral T-cells and increased susceptibility to infection/acute inflammation. In addition, we found that a single *N*-glycan antenna is the *sine-qua-non* condition needed to rescue homeostasis and T-cell developmental dynamics and peripheral phenotypes, due to a glycosylation pathway compensatory effect.

## RESULTS

### The glycocalyx landscape in human and murine T-cell development is dynamic and diverse

The complete glycocalyx composition and function of human and murine thymocytes is far from being completely deciphered. To gain insights into the composition and function of glycan structures at each T-cell developmental stage, an extensive lectin-based characterization of the human and mice thymic T-cell glycome was performed. A combination of different lectins recognizing specific *N*-glycan structures were used to assess the levels of relevant glycans along the *N*-glycosylation pathway (Fig. 1A). Thawed human thymocytes were used to perform the glycoprofiling of the main developmental T-cell populations (Fig. 1B, Supp. Fig. 1A). We found that human thymocyte populations display a unique and stage-selective glycocalyx composition that characterizes and distinguishes each developmental stage (Fig. 1C). Specifically, the complex branched *N*-glycans detected by L-PHA staining and the terminally α2,6 sialylated glycans detected by the SNA lectin, structures known to be involved in T-cell biology[7], displayed a clear distinctive profile along T-cell development (Fig. 1D-E). We observed a differential profile of complex branched *N*-glycans, containing a ß1,6-GlcNAc antenna (L-PHA binding), across human thymocyte populations (Fig. 1D), showing high representation of these *N*-glycans in DN cells, which decrease in ISP4, and finally increase in mature populations (Fig. 1F, top). Notably, between the DP pre-positive selection stage and both DP post-positive selection stage and mature SP cells, we found a significant increase in L-PHA reactivity (Fig. 1F, top), which reveals differential expression dynamics of complex branched *N*-glycans on thymocytes, when compared to peripheral cells[7]. By analyzing SNA reactivity we found an enrichment in α2,6-linked sialic acid in the mature SP populations (Fig. 1E), when compared to ISP4 and DP subsets (Fig. 1F, bottom). As shown by GNA binding, the proportion of less complex, high mannose *N*-glycans structures was low among all human thymocyte subsets, being more represented in mature SP populations (Supp. Fig. 1B, 1D top). The presence of elongated glycans (LEL binding) was higher in DN and ISP4 populations, reaching a peak in the latter (Supp. Fig. 1C, 1D bottom). These results show that human thymocytes display a unique and dynamic glycan signature that varies considerably during development.

**Fig. 1:**
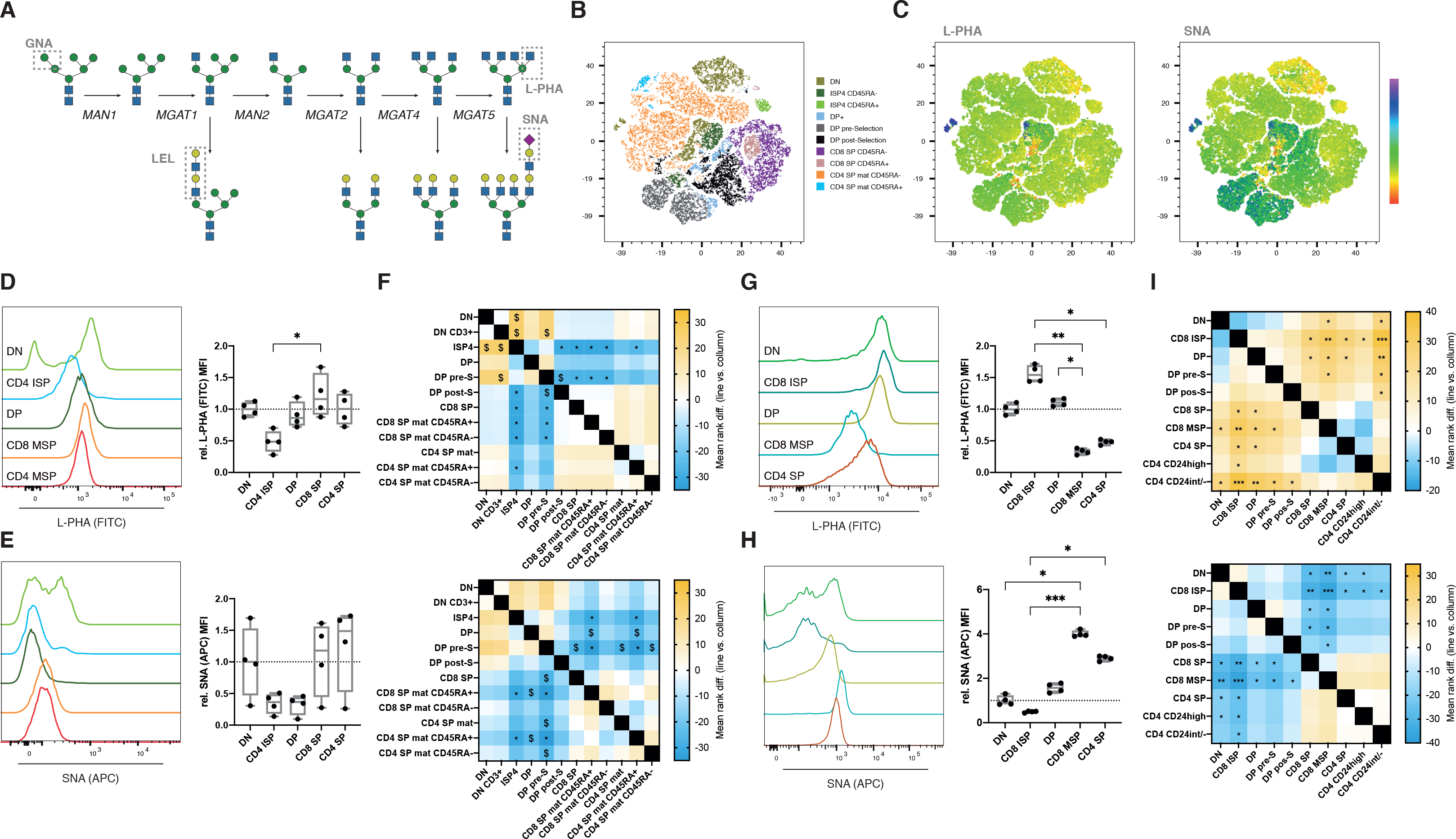
Human and murine T-cell developmental stages entail differential glycans profiles. **(A)**Schematic of the Golgi-located major steps of the *N*-glycosylation pathway, of GlcNAc antenna formation, elongation and termination. Boxes indicate the specific lectin binding glycans. **(B)**Unsupervised flow cytometry analysis of human thymocytes using tSNE, combining the data from 4 human thymus, with the main thymocyte populations annotated manually by the gating strategy defined (Supp. Fig. 1A). **(C)**Colormap for L-PHA (left) and SNA (right) binding levels in the same tSNE representation of **(B)**. Colorbar indicates the levels of binding. Histograms of L- PHA and of SNA binding in thymocyte subsets from human (**D** and **E**, respectively, N=4) and from mouse (**G** and **H**, respectively, N=4), with the right graphs depicting the Median Fluorescence Intensity (MFI) quantification and multiple comparisons. Heatmap showing the multiple comparison between populations, done as X (population in line) *vs*. Y (population in column). Data are mirrored across the diagonal. Color bar indicates the mean rank difference values. Kruskal-Wallis test, *q*-value $< 0.1, *< 0.05, **< 0.005, ***< 0.001.

Then, and to evaluate whether the glycan profile of murine thymocytes recapitulates the one displayed in humans, we glycophenotyped biologically equivalent murine thymocyte populations (Supp. Fig. 1E). In the mouse, the presence of complex branched *N*-glycans (L-PHA binding) was enriched in DN and DP cells, increasing in the ISP8 population (equivalent to the human ISP4). However, in mature SP populations, complex branched *N*-glycans decreased with developmental progression (Fig. 1G and 1I top). On the other hand, terminal α2,6-sialylation was found to be scarce in DN and ISP8 populations, reaching high levels in mature SP populations (Fig. 1H and 1I bottom). Moreover, high mannose *N*-glycans remained at low levels across all T- cell developmental stages (Supp. Fig. 1F and 1H top). Elongated glycans were found to have a developmental peak in ISP8 cells (Supp. Fig. 1G and 1H bottom). Interestingly, both human and mouse thymocytes shared major glycocalyx alterations displaying high levels of mature complex branched and elongated *N*-glycans in the ISP population that precedes the DP stage, and increased terminal α2,6-sialylation in mature subsets. Interestingly, the relative presence of complex branched *N*-glycans in mature SP (increase/maintenance in human and decrease in mouse) may underlie differences between organisms regarding thymic egress and peripheral naïve T-cell properties.

Taken together, this analysis of human and murine thymocytes suggests the existence of a developmentally-regulated glycome, which supports the existence of functional effects of glycans in the regulation of T-cell development.

### Lack of GlcNAc-branching *N*-glycans impairs T-cell development, which is rescued by a compensatory mono-antennary *N*-glycan structure

The relative abundance of mature complex branched *N*-glycans in DN and ISP developmental stages (Fig. 1) and their ability to control TCR thresholds[13] prompted us to investigate their functional relevance in the context of T-cell development. We hypothesized that complex *N*-glycan structures might have a prominent role during pre-mature TCR selection. To test this, we have genetically modified the *N*-glycosylation pathway in murine models, at early DN stages, using a *Rag1*^Cre^ strain crossed with *Mgat1*^fl/fl^ or *Mgat2*^fl/fl^ ones, thus eliminating these two glycogenes as early as in the DN2 developmental stage. The mouse model with the *Rag1*^Cre^- mediated knockout of *Mgat1* (*Mgat1* cKO) display a complete truncation of complex branched *N*- glycans, only expressing high-mannose *N*-glycans, whereas the conditional knockout of *Mgat2* (*Mgat2* cKO) exhibits a partial truncation of complex branched *N*-glycans, generating hybrid-type *N*-glycan structures with only one GlcNAc antenna which can be further extended. We then set out to evaluate alterations of the most prevalent thymocyte populations in both glycoengineered models (Fig. 2A and 2I).

**Fig. 2:**
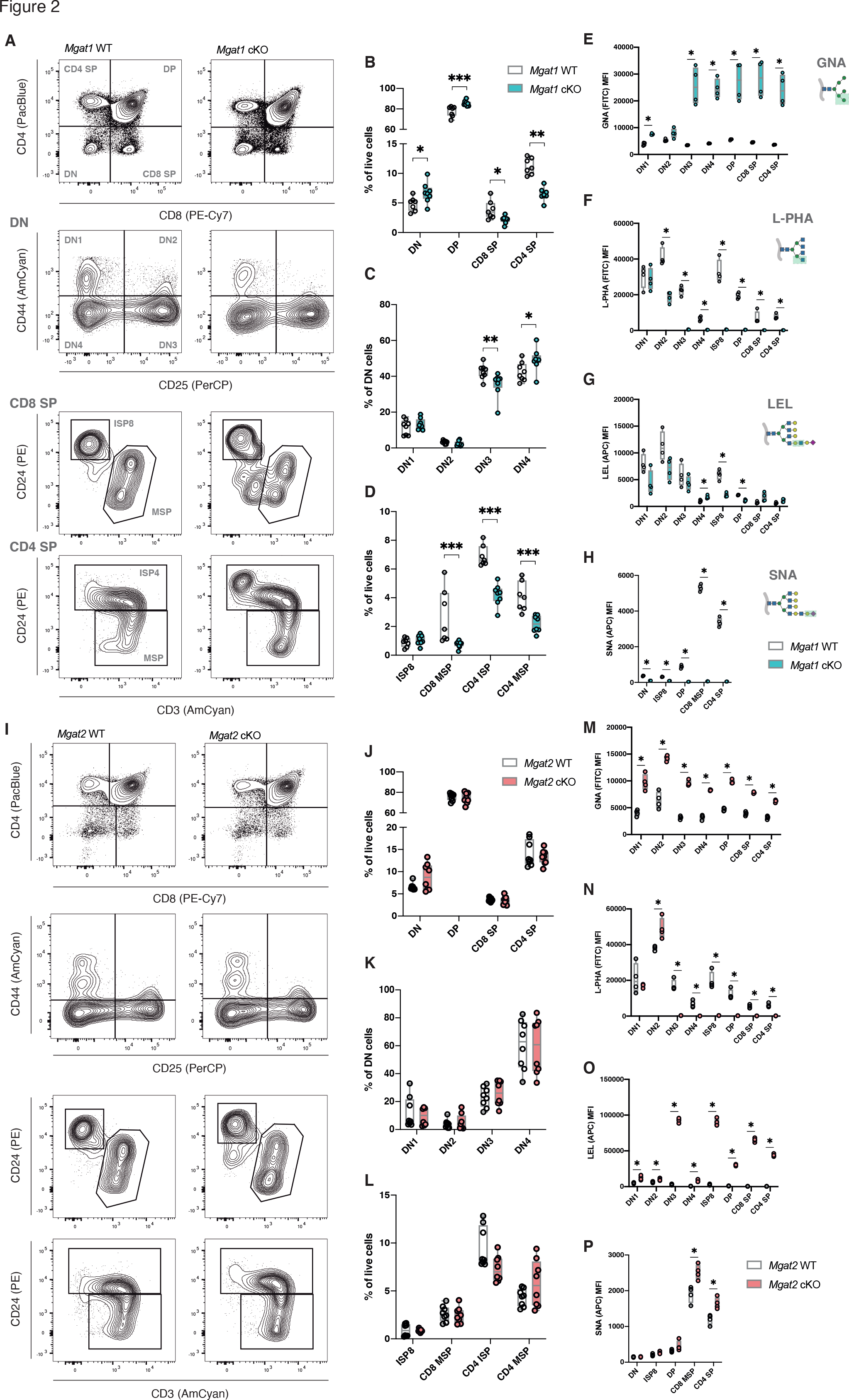
Impairment of T-cell development in high-mannose restricted thymocytes. Thymocyte population discrimination in *Mgat1*WT (N = 4 to 8) and *Mgat1* cKO (N = 4 to 8) and in *Mgat2*WT (N = 4 to 8) and *Mgat2* cKO (N = 4 to 8) mice (**A** and **I**, respectively). Frequencies of DN (CD4- CD8-), DP (CD4+CD8+), CD8 SP (CD4-CD8+) and CD4 SP (CD4+CD8-) thymocyte subsets, in both models (**B** and **J**). Frequencies of DN1 (CD44+CD25-), DN2 (CD44+CD25+), DN3 (CD44- CD25+) and DN4 (CD44-CD25-) subsets, within the total DN population, in both models (**C** and **K**). CD8 SP and CD4 SP subset frequencies: ISP8 (CD8+CD24+CD3-), CD8 MSP (CD8+CD24int/-CD3+), CD4 ISP (CD4+CD24hiCD3lo/hi) and CD4 MSP (CD4+CD24loCD3hi), in both models (**D** and **L**). (**E** and **M**)GNA, (**F** and **N**) L-PHA, (**G** and **O**) LEL and (**H** and **P**) SNA binding levels (MFI) in the thymocyte subsets. Each dot represents one mouse. Mann-Whitney t-test, *p*-value *< 0.05, **< 0.005 and ***< 0.001.

Notably, *Mgat1* cKO mice display a significant increase in fraction of DN and DP thymocytes populations, which is accompanied by a significant decrease in CD8 SP and CD4 SP populations (Fig. 2B). Within the DN compartment, complete truncation of complex branched *N*- glycans resulted in a significant drop of DN2 and DN3 thymocytes and an increased frequency of DN4 thymocytes (Fig. 2C), suggesting a significant role of these carbohydrate structures in ß- selection. Moreover, a significant decrease in CD8 mature SP cells, CD4 immature (CD24^hi^, post- selection and pre-egress) and mature (CD24^lo^) SP cells was observed (Fig. 2D), which may reflect increased DP negative selection. However, no differences were observed in the frequency of ISP8 thymocytes between *Mgat1* proficient and deficient models (Fig. 2D). The total number of thymocytes were not different when *Mgat1* pro- and deficient thymi were compared, however with some significant population specific alterations (Supp. Fig. 2A). Since the absence of complex branched *N*-glycans may influence expression of cell surface glyco-receptors, we evaluated the expression levels for the CD4 and CD8 co-receptors, relevant for DP selection/lineage commitment, and found a significant decrease of both CD4 and CD8 SP co-receptors in the respective SP populations (Supp. Fig. 2B-C).

We then evaluated the impact of *Mgat1* deletion on glycocalyx composition of thymocytes. As expected, we observed a sticking increase in high mannose *N*-glycans (GNA binding) in all developmental stages beyond DN2 (Fig. 2E, Supp. Fig. 2I top left). Accordingly, interruption of the *N*-glycosylation pathway led to a complete absence of complex branched *N*-glycans, as shown by L-PHA staining, in all developmental stages beyond DN2 (Fig. 2F, Supp. Fig. 2I top right). Elongated glycans (LEL binding) were unaltered in DN cells, suggesting their presence in *O*- glycans, but were severely decreased in ISP8 and DP thymocyte populations (Fig. 2G, Supp. Fig. 2I bottom left). Loss of complex branched *N*-glycans was also accompanied by ablation of terminal α2,6-sialic acid, as determined by SNA probing, in thymocyte’s glycocalyces (Fig. 2H, Supp. Fig. 2I bottom right).

These results led us to investigate whether the presence of a single GlcNAc-antenna in *N*- glycans would be sufficient to ensure normal developmental processes. Interestingly, we observed that the frequencies of thymocyte populations in *Mgat2* cKO mice were fully mitigated and compensated, when compared to their wild-type counterparts (Fig. 2I and 2L). Thus, the presence of hybrid *N*-glycans, containing the first GlcNAc antenna generated by GnT-I (encoded by *Mgat1*), was sufficient and a *sine-qua-non* condition to guarantee normal thymic T-cell development. Notably, the total number of thymocytes of *Mgat2* cKO mice was unaltered, however with some significant population specific alterations (Supp. Fig. 2E). Moreover, we observed a decreased expression of the CD4 and CD8 co-receptor surface expression in their respective SP subsets upon *Mgat2* deficiency (Supp. Fig. 2F-G).

The *N*-glycome that enables homeostatic development is characterized by a significant increase of higher levels of mannose residues (GNA binding), present in hybrid *N*-glycans (Fig. 2M, Supp. Fig. 2J top right) and absence of the ß1,6-GlcNAc antenna (L-PHA binding) in the developmental stages past DN2 (Fig. 2N, Supp. Fig. 2J top right). Interestingly, we also found a remarkable increase of elongated poly-LacNAc structures (LEL binding) from the DN3 stage (Fig. 2O, Supp. Fig. 2J bottom left), along with a significant increase of terminal α2,6-sialylation that was evident in mature CD8 and CD4 SP populations (Fig. 2P, Supp. Fig. 2J bottom right). Thus, an unique GlcNAc-antenna is found to be hyper-elongated, similar to what was described in *Mgat2* deficient mature T-cells[17], generating a compensatory *N*-glycan structure, that is sufficient to guarantee normal T-cell development, that is prevented when mature complex branched *N*-glycans are absent (in a *Mgat*1 deficiency scenario). Interestingly, we found that compensatory *N*-glycan hyper-elongation also restored binding of endogenous galectin-3 (Gal-3) and galectin-1 (Gal-1), β-galactoside-binding lectins, to the surface of DP thymocytes (Supp. Fig. 2H), which were not observed in *Mgat1-*deficient thymocytes (Supp. Fig. 2D).

Collectively these data suggest that the loss of *N*-glycan complexity, in the *Mgat1* cKO model, has a severe influence on T-cell development, precluding the selection of peripheral T-cells. This effect could be rescued in the *Mgat2* cKO model, suggesting that one GlcNAc antenna *N*-glycan is sufficient to compensate the *N*-glycosylation pathway and guarantee normal T-cell development.

### Efficient ß-selection requires *N*-glycan branching

Given the alterations in the frequency of DN cells imposed by *Mgat1* deficiency, particularly the decrease of DN3 and the increase in DN4 subpopulations (Fig. 2), we sought to investigate the impact of complex branched *N*-glycans in ß-selection. This key developmental checkpoint translates into the functionality of the newly generated ß-chain TCRs, a product of *Tcrb* gene rearrangements[1, 2]. ß-selected DN3 cells begin to express pre-TCR complexes, detected by the expression of intracellular (ic)TCRß chain[18]. To evaluate the role of *N*-glycan branching in ß-selection, we first determined the frequency of DN3 cells bearing icTCRß. We found in the *Mgat1* cKO model, a significant decrease of icTCRß+ DN3 cells but not DN4 cells (Fig. 3A-B), suggesting that clonal expansion of ß-selected thymocytes was not compromised, but rather their generation. Moreover, in the absence of *Mgat1*, a decrease in CD25 surface expression was detected in total DN3 cells (Fig. 3C), suggesting ß-selection defects[19]. Pre-TCR signaling is responsible for induction of DN3 proliferation and DN4 transition, and can be detected using the proxy CD5 expression[20]. In the *Mgat1* cKO model, we found a decrease of CD5 expression in DN3, but not in DN4 cells (Fig. 3D). Thus, disruption of complex branched *N*-glycans interrupts generation of ß-selected thymocytes, but does not alter their clonal expansion. Moreover, as the disruption of ß1,6-GlcNAc branching *N*-glycans promotes mature TCR receptor clustering and T- cell hyperactivation[7, 15], the clustering of pre-TCR complexes could be playing a role. As IL-7 signaling is one of the key regulators of ß-selection[1, 2] we evaluated the IL-7Rα (CD127) expression in DN3 cells and found a decrease in *Mgat1* cKO thymocytes (Fig. 3E).

**Fig. 3:**
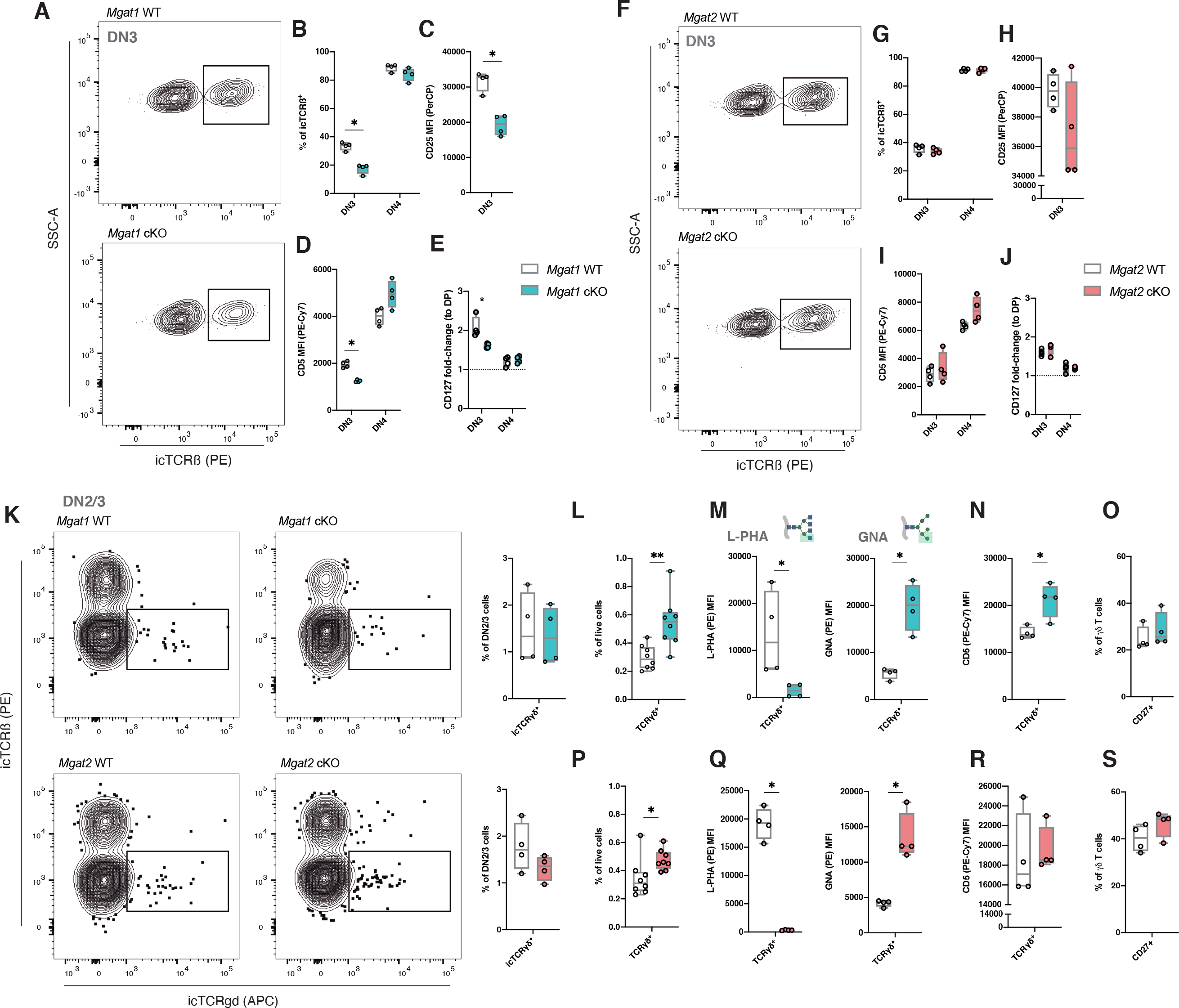
Lack of complex branched N-glycans impairs ß-selection and favors determination of γδ T-cells. Identification of ß-selected (icTCRß+) DN3 cells, in *Mgat1*WT (N = 4) and *Mgat1* cKO (N = 4) and in *Mgat2*WT (N = 4) and *Mgat2* cKO (N = 4) mice (**A** and **F**, respectively). Frequency of icTCRß+ cells in DN3 and DN4 thymocytes, in both models (**B** and **G**). Quantification of CD25 surface levels (MFI) in the DN3 subset, in both models (**C** and **H**). Quantification of CD5 surface levels (MFI) in DN3 and DN4 thymocytes, in both models (**D** and **I**). Quantification of CD127 surface levels (MFI), normalized for each mouse to the DP population, in DN3 and DN4 thymocytes, in both models (**E** and **J**). Identification of icTCR*γδ*+ cells within the DN2 and DN3 populations, and frequency quantification, for *Mgat1*WT (N = 4), *Mgat1* cKO (N = 4), *Mgat2*WT (N = 4) and *Mgat2* cKO (N = 4) mice (**K**). Frequency of total TCR*γδ*+ cells, in both models (**L** and **P**). L-PHA (left) and GNA (right) binding levels, in both models (**M** and **Q**). CD5 surface levels (MFI) of TCR*γδ*+ cells, in both models (**N** and **R**). CD27 surface levels (MFI) of TCR*γδ*+ cells, in both models (**O** and **S**). Each dot represents one mouse. Mann-Whitney t-test, p-value *< 0.05, **< 0.005 and ***< 0.001.

In accordance with previous results, the presence of the mono-antennary *N*-glycans on *Mgat2* cKO mice, was found to be sufficient to compensate and reestablish the ß-selection process (Fig. 3E-J). Thus, complex branched *N*-glycans are essential structures that control ß-selection and pre-TCR signaling.

### Disruption of *N*-glycan branching favors positive selection of γδ T-cell development

T-cell commitment towards the *γδ* T-cell lineage occurs early at the DN2 and DN3 stages, where the *Tcrg* and *Tcrd* may also be targeted for genetic rearrangements[1, 2]. In spite of considerable progress, the precise mechanisms underlying lineage commitment have not been fully clarified[21]. Since the absence of *Mgat1* leads to deficient DN3 ß-selection (Fig. 3A-B), we analyzed the presence of DN2/3 cells bearing intracellular (ic)TCR*γδ*. However, we found no differences in either *Mgat1-* and *Mgat2-*deficient thymocytes (Fig. 3K). Furthermore, an increase in the proportion of mature *γδ* T-cells was observed in both glycoengineered models (Fig. 3L and

3P). These cells were shown to display absence of complex branched *N*-glycans (L-PHA binding) and expression of mannosylated *N*-glycans (GNA binding), in both models, as expected (Fig. 3M and 3Q). When evaluated for TCR-mediated activation, using the CD5 marker, we observed increased CD5 expression in *γδ* T-cells of *Mgat1* cKO (Fig. 3N), but not in *Mgat2* deficient mice (Fig. 3R), suggesting higher TCR signaling in *Mgat1* deficient *γδ* T-cells, which has been associated with the promotion of this T-cell subset development[21]. As IFN-producing *γδ* T-cells derive from CD27^+^ thymic progenitors[22], we evaluated the frequency of CD27^+^*γδ* T-cells, but found no differences in both models (Fig. 3O and 3S).

Our results suggest that complex branched *N*-glycans are major determinants for *αβ* and *γδ* T lineage discrimination.

### Complex branched *N*-glycans are major regulators of TCR repertoire diversity

Previous evidences showed that *N*-glycans regulate positive and negative DP selection, in a *Lck*^Cre/+^*Mgat1*^fl/fl^ background, where ∼70% DP cells were defective in complex-branched *N*- glycans.^13^ In our *Mgat1*cKO model, in which all DP cells are deficient in complex-branched *N*- glycans, the DP selection effect was validated. To investigate the impact of complex *N*-glycans in TCR-selection, we analyzed the differential expression of the TCRß chain and CD69, to identify the thymocyte populations undergoing selection^23^ (Fig.4A and 4D). Interestingly, *Mgat1* deficiency caused a significant frequency increase of TCRß^-^CD69^-^ cells (Fig.4B), that had higher levels of apoptosis, seen by annexin V staining (Fig.4C), indicating increased death by neglect. Accordingly, a decrease in positively selected cells, TCRß^hi^CD69^hi^, was observed (Fig.4B). Finally, post-selection thymocytes, TCRß^hi^CD69^-^ were reduced in *Mgat1*cKO mice (Fig.4B), suggesting more pronounced negative selection. Again, these differences were compensated in the *Mgat2*cKO mice, as the populations of DP selection as well as apoptosis levels were similar to their control counterparts (Fig.4D-F).

**Fig. 4:**
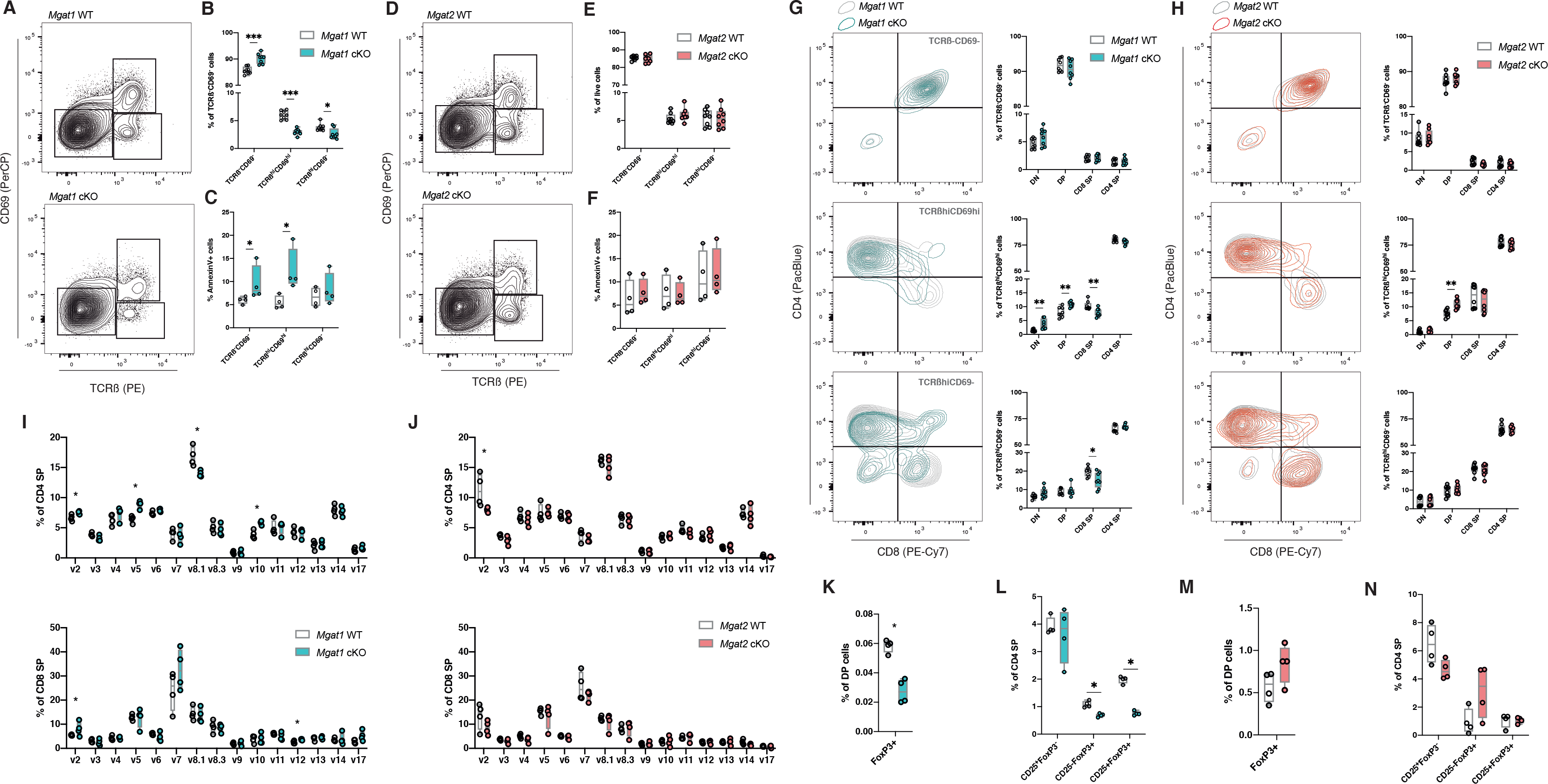
TCR-selection is impaired in high-mannose restricted *N*-glycome, with severe effects in TCR repertoire diversity and thymic Treg generation. (A) and (D) Identification of pre- selection (TCRß^-/lo^CD69^-^), post-positive selection (TCRß^int/hi^CD69^int/hi^) and post-negative selection (TCRß^hi^CD69^-^) thymocytes in *Mgat1*WT (N = 8) and *Mgat1* cKO (N = 8), and *Mgat2*WT (N = 8) and *Mgat2* cKO (N = 8) mice, respectively. (B) and (E) Quantification of the frequencies of the populations identified in (A) and (D). (C) and (F) Quantification of Annexin V+ cells within the thymocyte populations in *Mgat1*WT (N = 4) and *Mgat1* cKO (N = 4), and *Mgat2*WT (N = 4) and *Mgat2* cKO (N = 4) mice, respectively. (G) and (H) Distribution of each selection subset according to its CD4 and CD8 expression levels, and quantification in *Mgat1*WT and cKO, and *Mgat2*WT and cKO mice, respectively. (I) and (J) Screen of TCRvß+ expressing cells, within mature CD4 SP (top) and CD8 SP (bottom) in both models. (K) and (M) Quantification of the frequency of FoxP3+ cells in the DP population, in *Mgat1*WT and cKO, and *Mgat2*WT and cKO mice, respectively. (L) and (N) Detection of Treg thymic precursors, by the expression of CD25 and FoxP3, and population quantification, in *Mgat1*WT and cKO, and *Mgat2*WT and cKO mice, respectively. Each dot represents one mouse. Mann-Whitney t-test, p-value *< 0.05, **< 0.005 and ***< 0.001.

We then analyzed the CD8/CD4 lineage commitment, occurring during TCR-selection, by analyzing the relative distributions of the three populations described above, according to CD8/CD4 expression (Fig.4G-H). Interestingly, positively selected cells were differentially distributed according to their CD8 and CD4 surface expression in both models (Fig.4Gmiddle). These results indicate a role for *N*-glycans on CD4/CD8 lineage commitment.

One of the most important events occurring during TCR-selection is the generation of a diverse TCR-repertoire.^23^ Given the impact of *N*-glycans in TCR-selection, we evaluated whether selection defects observed in *Mgat1*cKO mice could generate impaired TCR diversity. For this, we screened for TCRß chain expression in the mature SP subsets, using a panel of monoclonal antibodies. Notably, in *Mgat1*cKO mice we found a significant impact on the diversity of the TCRß-variants-expressing cells in mature SP subsets (Fig.4I). However, no major differences on TCR-repertoire were observed for *Mgat2*cKO thymocytes(Fig.4J). We cannot exclude that the defects on ß-selection of *Mgat1cKO* mice may contribute to this phenotype and further studies on TCR-repertoire diversity analysis using next-generation sequencing would provide further insights.

These results indicate the central role of complex-branched *N*-glycan structures in DP lineage commitment and TCR-repertoire diversity.

### Deficiency in complex branched *N*-glycans hampers regulatory T-cell development

The ability of complex branched *N*-glycans to control the thresholds of TCR positive and negative selection raised the question as whether these structures could influence thymic development of T regulatory cells (Tregs). In fact, naturally-occurring Tregs (nTregs) are generated in the thymus, comprising a set of CD4 SP cells with high levels of activation, close to the threshold of negative selection[24]. We have shown that negative selection is promoted in *Mgat1* cKO mice (Fig. 4C) and we argued that Treg generation would be impaired. The initial precursors of Tregs are DP cells which show considerable expression of the FoxP3 transcription factor. Interestingly, this population is decreased in an *Mgat1* deficient scenario (Fig. 4K), whereas no differences were found for *Mgat2* deficiency (Fig. 4M). As mature Tregs may arise from CD4^+^CD25^+^FOXP3^+^ and CD4^+^CD25^-^FOXP3^lo^ progenitors, we evaluated the presence of these two subsets in the thymic compartment. We detected lower frequency of both populations in *Mgat1* cKO mice (Fig. 4L), and no differences were found in *Mgat2* cKO ones (Fig. 4N). Thus, our results highlight an impact of complex branched *N*-glycans on thymic Treg generation and this effect can be rescued in the presence of hyper-elongated hybrid *N*-glycans.

### Lack of complex branched *N*-glycans impairs thymic egress of mature thymocytes

The final step of T-cell development is thymic egress, where thymocytes that surmounted negative selection upregulate CD62L expression and leave the thymus[25]. The presence of mature and selected CD4 and CD8 SP thymocytes in both murine models was assessed and the relative expression of CD3 and CD69 was used to detect post-selection cells (Fig. 5A-B). A remarkable decrease in the frequency of CD62L^+^ cells was observed in post-selection CD4 SP in the *Mgat1* cKO mice, that was fully compensated in *Mgat2* cKO ones (Fig. 5A). However, no differences in the CD8 SP subset (Fig. 5B) were detected, although a trend toward a decrease in CD8 SP subset could be observed in the absence of branched *N*-glycans. Thus, thymic egress is markedly compromised when complex *N*-glycans are absent.

**Fig. 5:**
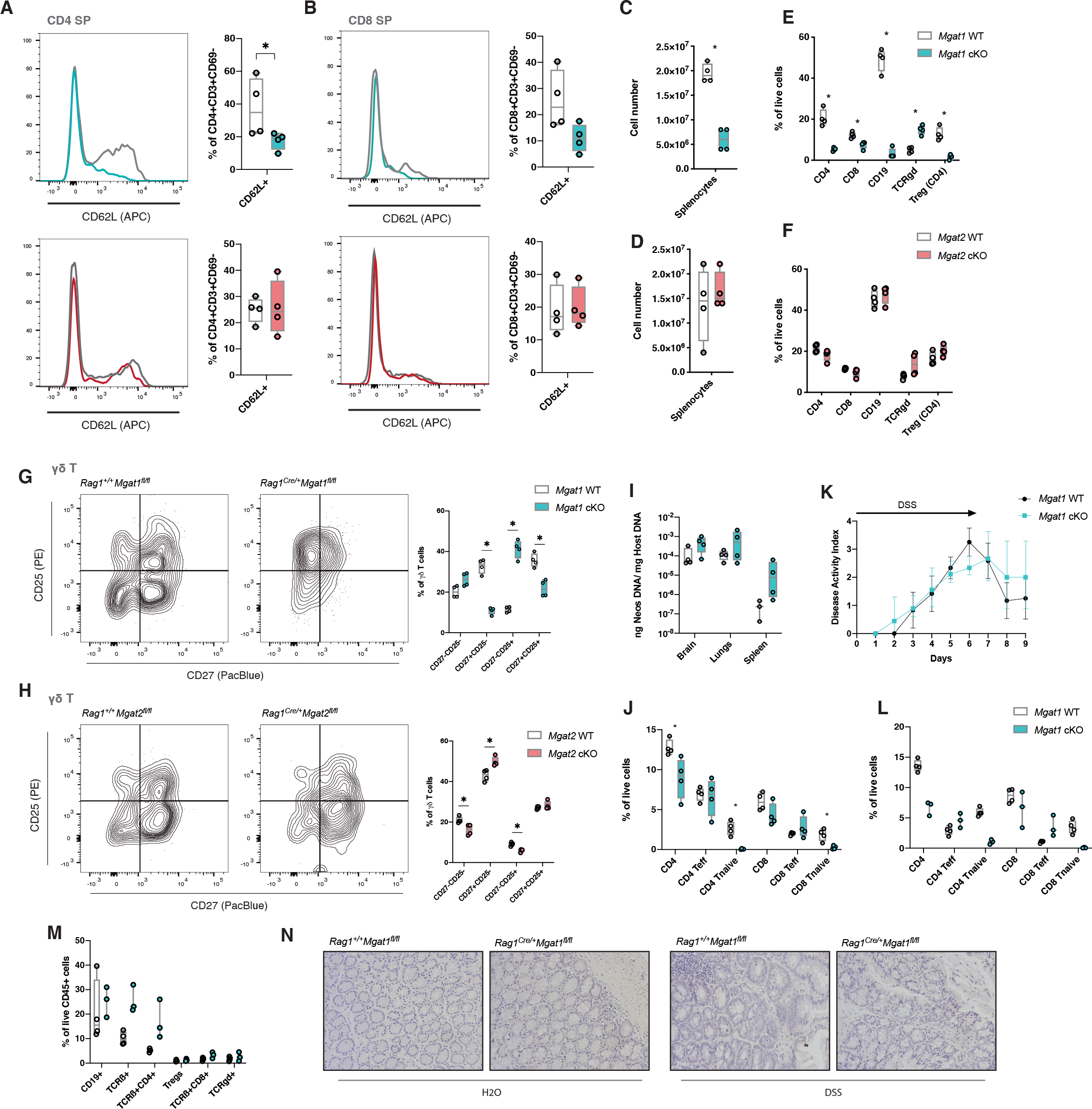
Impairments on T-cell development in the absence of *Mgat1* have T-cell egress and peripheral consequences. **(A)** and **(B)** Identification of CD62L^+^ cells in mature CD4 SP and CD8 SP, in *Mgat1*WT (N = 4) and *Mgat1* cKO (N = 4), and *Mgat2*WT (N = 4) and *Mgat2* cKO (N = 4) mice, respectively. **(C)** and **(D)** Total splenocyte cell numbers in *Mgat1*WT and cKO, and *Mgat2*WT and cKO mice, respectively. **(E)** and **(F)** Quantification of CD4 T, CD8 T, CD19+ (B cells), *γδ* T and Tregs (within CD4 T) immune cell splenic frequencies, in *Mgat1*WT and cKO, and *Mgat2*WT and cKO mice, respectively. **(G)** and **(H)** Peripheral *γδ* T-cell activation and differentiation analysis, by the expression of surface CD25 and CD27, and quantification, in *Mgat1*WT and cKO, and *Mgat2*WT and cKO mice, respectively. **(I)** *N. caninum* organ colonization determination, through the quantification of total parasite DNA in 1 mg of total host DNA, in *Mgat1*WT (N = 3 - 4) and *Mgat1* cKO (N = 4). **(J)**T-cell frequencies of the spleens of infected mice. **(K)**Disease Activity Index (DAI) scores for the DSS-induced colitis in *Mgat1*WT (N = 4) and *Mgat1* cKO (N = 3). **(L)**T-cell frequencies of the spleens and **(M)** of the colons of the mice. **(O)** Representative histological analysis of both genotypes, at the final day of the experiment, for both H2O and DSS-water fed animals. Each dot represents one mouse. Mann- Whitney t-test, p-value *< 0.05, **< 0.005 and ***< 0.001.

### The absence of complex branched *N*-glycans in thymocytes impairs peripheral T-cell homeostasis and immune response

Given the number of defects observed in T-cell development upon complex branched *N*- glycan deficiency, we further explored the biological consequences of these effects within the peripheral compartment in both models. We have observed a drastic reduction in total splenocyte numbers in *Mgat1* cKO mice (Fig. 5C), with no impact observed in *Mgat2* cKO ones (Fig. 5D). In terms of frequencies of immune populations, we observed a close to absent expression of both T and B cell subsets in the spleens of *Mgat1* cKO mice (Fig. 5E), comparing with *Mgat2* deficient ones that showed a preserved expression of these immune cell populations in the spleen similar to controls (Fig. 5F). This immunoprofile of *Mgat1* cKO mice is compatible with a primary immunodeficient phenotype. We then evaluated the TCRvß repertoire in splenic T-cells and found critical differences in *Mgat1* deficient CD4 and CD8 T-cells, with no alterations in the *Mgat2* cKO model (Supp. Fig. 3A-B). Strikingly, we found a considerable expansion of the *γδ* T-cell compartment in the spleens of *Mgat1* cKO mice (Fig. 5E), mimicking a genetic model of *αβ*T-cell depletion. Moreover, and using the activation marker CD25 and the differentiation marker CD27, we found substantial alterations in homeostatic *γδ* T-cell differentiation. Lack of branched *N*- glycans increased the activation status of *γδ* T-cells, but impaired their differentiation status as revealed by a decrease of CD27^+^CD25^+^ T-cells (Fig. 5G). Curiously, and despite no significant differences on total *γδ* T frequencies on *Mgat2* cKO mice (Fig. 5F), we did observe alterations in *γδ* T differentiation (Fig. 5H). To our knowledge, this is the first report demonstrating the impact of branching *N*-glycosylation pathway in peripheral *γδ* T-cell differentiation and activity.

We next explored the pathophysiologic consequences in terms of disease susceptibility of these phenotypic changes. Both *Mgat1* cKO and *Mgat2* cKO mice were challenged with the obligate intracellular protozoan *N. caninum*, as resistance to this parasite was previously shown to be highly T-cell-dependent[26]. Our results showed that *Mgat1* cKO mice exhibited an increased parasite colonization in key organs analyzed, suggesting a tendency for an increased susceptibility to infection, when compared to their wild-type counterparts (Fig. 5I), that was not observed in *Mgat2* cKO mice (Supp. Fig. 3C). Since *N. caninum*-related immune responses are highly dependent on an intact Th1 response[26], we further analyzed the immunophenotype of the splenocytes of infected mice, and observed an expansion in both CD4^+^ and CD8^+^ T-cell frequencies in *Mgat1* cKO mice, when compared to the steady state ones (Fig. 5J), that presented a higher frequency CD69+ cells in CD4^+^ T-cells, upon infection, which agrees with the role of complex *N*-glycans in TCR clustering (Supplemental Fig. 3D). Moreover, we detected negligible levels of naïve T-cells (CD62L^+^CD44^-^) in the spleens of these mice (Fig. 5J). These results support an increased pro-inflammatory response in *Mgat1* cKO mice that, together with lowered thresholds for T-cell activation, may explain the increased susceptibility to infection.

To gain further insights on the impact of complex branched *N*-glycans in T-cell development and on susceptibility to inflammation, we induced experimental colitis in both models, using dextran sulfate sodium (DSS), given our previous findings on the critical role of T- cell glycosylation in Inflammatory Bowel Disease[7, 14]. We found a trend toward increased susceptibility of *Mgat1* cKO mice to colitis (Fig. 5K, and Supp. Fig. 3E), as compared with *Mgat2* cKO mice and controls (Supp. Fig. 3F). Moreover, in the *Mgat1* cKO spleens we found a significant representation of effector T-cell subsets (CD62L^-^CD44^+^), within the CD4 and CD8 T-cell compartments (Fig. 5L). Particularly, in the colon of these mice, we found an increased proportion of T-cells and a higher immune infiltrate (Fig. 5M-N), in agreement with a pro-inflammatory response and decreased activation thresholds of T-cells[7]. Interestingly, in the H2O controls, higher immune infiltrates are seen in the *Mgat1* cKO mice (Fig. 5N). Altogether, our results show that an early thymocyte deficiency of complex branched *N*-glycans, has profound central and peripheral immune consequences and confers increased susceptibility to infection and immune-mediated pathology. Hyper-elongation of *N*-glycans with GlcNAc antennae efficiently rescues this T-cell phenotype and prevents higher susceptibility to disease development.

## DISCUSSION

T-cell development is a key biological process that ensures a functional and diverse TCR repertoire of T-cells, that enables robust mechanisms of immune protection and tolerance. The discovery of cellular developmental mediators of thymocyte maturation has deserved attention since the 60’s, and have shaped modern immunology[27]. However, in spite of considerable progress, the function of cell-specific *N*-glycans in early T-cell commitment, repertoire and selection, and their relevance in T-cell-mediated immunopathology, has been largely unexplored. Here we present a comprehensive characterization of the glycocalyx composition of both human and murine thymocytes, demonstrating the existence of an unique glycosylation signature that distinguishes developmental stages. In both organisms, in homeostatic conditions, thymocytes display high levels of complex branched *N*-glycans and an upregulation of terminal α2,6- silalylation in mature SP populations. In fact, this analysis of the glycome highlighted high *N*- glycan complexity across different T-cell developmental stages, namely the high levels of poly-LacNAc structures in immature subsets, elevation of terminal α2,6-silalylation in mature SP populations, both accompanied by a considerable presence of complex ß1,6-GlcNAc branching. Moreover, the overall low levels of high mannose structures, suggests *N*-glycan complexity as a *sine-qua-non* feature for a proper and functional T-cell development.

In order to model the expression of mature complex *N*-glycans in thymocytes, we generated unique glycoengineered mouse models with conditional ablation of branched *N*-glycans structures in T-cells, through the *Rag1*^Cre^-mediated disruption of the *Mgat1* and *Mgat2* in DN thymocytes. A deficiency in the expression of complex branched *N*-glycans, modelled in *Mgat1* cKO mice, resulted in increased percentages of DN cells relative to total thymocytes that were associated with an impairment of ß-selection. In total DN3 cells, we showed a decreased pre-TCR signaling in the absence of *Mgat1*, that was restored in DN4, which can be due to pre-TCR clustering, as seen in mature TCR’s, upon the deficiency of complex branched *N*-glycans[13–16]. Moreover, decreased expression of the IL-7R*α* may contribute to impaired ß-selection, and DN3 survival. The decreased frequency of ß-selected cells appears to be the result of a blockade in *α*ß/*γδ* lineage fate commitment, as demonstrated by an increased *γδ* T-cell population (resembling *Tcrb*^-/-^ or *Tcra*^-/-^ mice[30]) upon the deficiency of *Mgat1*, highlighting the relevance of *N*-glycans as master regulators of lineage development and commitment. Moreover, as *N*-glycans control *α*ßTCR thresholds, the same regulatory effect might be extrapolated to *γδ*TCRs, promoting sustained signaling to *γδ* T-cells (as shown in the *Mgat1* deficient model), and favoring development of this T-cell subset[21]. To our knowledge, this is the first report on the role of *N*-glycans in *γδ* T-cell development, suggesting the critical role of these complex carbohydrate structures in this T-cell subset.

Elegant studies on the contribution of *N*-glycans in TCR selection showed its impact in the regulation of the thresholds for positive and negative selection[13]. Our work, targeting *Mgat1* at early stages of T-cell development (*Rag1*^Cre^), has illuminated the contribution of *N*-glycans in several critical T-cell developmental processes, including lineage commitment, TCR repertoire determination, thymic Treg generation, *γδ* T-cell development and thymic egress. DP thymocytes are bi-potential progenitors for the CD4+ and CD8+ T-cell lineages, during positive selection[23].

We found that the lack of complex *N*-glycans in cells that have been positively selected was associated with an increased percentage of cells in the DN and DP quadrants, indicating enhanced co-receptor internalization, as previously observed in other settings[13], which altered the fate of DP cells to the CD4 SP and CD8 SP lineages. Importantly, our results further identify an additional and novel role of *N*-glycans in TCR selection, demonstrating that a deficiency in complex branched *N*-glycans significantly alters the diversity of the TCRvß variants in mature SP cells. Moreover, we found that thymic Treg generation is also compromised in the absence of complex branched *N*-glycan structures. Impairment in the generation of nTregs could be associated with reduction of the TCR affinity threshold for negative selection imposed by the absence of *N*-glycan branching[13]. Finally, we found a deficiency in CD62L-expressing mature CD4 SP thymocytes in *Mgat1* cKO’s, suggesting an impairment of egression. Altogether, our results demonstrated that thymocyte glycocalyces are developmentally and dynamically regulated, being complex and elongated branched *N*-glycan structures essential to ensure proper transitions between developmental stages and for regulating central selection and commitment processes.

Interestingly, in this study, we further demonstrated that GlcNAc-branching of *N*-glycans is a *sine-qua-non* condition for appropriate T-cell development as highlighted by the fact that *Mgat2* cKO mice, proficient in synthesizing in mono-antennary *N*-glycan structures in T-cells, do not display major developmental defects, exhibiting a normal ß-selection, production of diverse TCR repertoires, normal generation of Tregs and thymic egress, as well as *γδ* T-cell development.

This suggests that hyper-elongated mono-antennary *N*-glycans are key glycan determinants essential in T-cell development and able to rescue the majority of developmental processes markedly impaired in *Mgat1* deficient mice. The synthesis of a mono-antennary *N*-glycan structure appears to create a compensatory mechanism that allows glycan elongation and terminal sialylation, essential to feed a proper T-cell selection program. Interestingly, these glycosylation compensatory pathways have been described in other pathophysiologic settings[17, 33].

The phenotypic impact of such a drastic impairment on T-cell development upon absence of complex branched *N*-glycan structures was demonstrated in *Mgat1* cKO mice. We demonstrated that a central deficiency of complex branched *N*-glycans on T-cells results in remarkable reduced lymphocyte populations at the periphery, namely *αβ*T and B cells, resembling primary immunodeficient disorders. The TCRvß reduced variability in thymic SP cells was observed to be conserved in splenic T-cells, in the *Mgat1* deficient model. This immunodeficient phenotype was further validated by the increased susceptibility to infection and intestinal inflammation, again fully compensated by the presence of one GlcNAc-antenna. In addition, we found that *γδ* T-cells were able to survive and escape to the periphery upon branched *N*-glycans deficiency, pinpointing these cells as key players in the increased susceptibility to infection and inflammation, a concept that deserves future exploration.

Taken together, this study unveils the regulatory power of branched *N*-glycans in T-cell development, highlighting their role as key determinants for immunity and as essential players in the control of ß-selection, Treg generation, TCR repertoire diversity and *γδ* T-cell development. Our findings constitute novel features of thymocyte “identity” and function, by revealing branched *N*-glycans as essential structures of T-cells with implications in determining the susceptibility to the development of major diseases, such as infection and inflammation.

## MATERIALS AND METHODS

### Study design

The goal of this study was to identify differential glycoprofiles of human and murine thymocytes, and whether the glycoengineering of murine ones would result in alterations in normal T-cell development and response. We generated murine conditional knockout models for Mgat1 and Mgat2, with a Rag1Cre line. Using lectin- or antibody-based flow cytometry, we detected cellular subsets’ glycoprofiles and alterations in homeostatic T-cell development, through phenotyping thymocyte populations. To assess the contribuition of the Mgat1 and Mgat2 defficiency in T-cell peripheral responses we used a model of infection and acute inflammation induction. All experiments/analysis were performed two or more times.

### Experimental model and subject details Animals

Mice were housed at the animal facility of the Institute for Research and Innovation in Health of the University of Porto (i3S, Porto, Portugal). C57BL/6 wild-type (WT) mice were acquired from the Jackson laboratory. Mice containing floxed *Mgat1* (MSR Cat# JAX:006891, RRID:IMSR_JAX:006891) and *Mgat2* (IMSR Cat# JAX:006892, RRID:IMSR_JAX:006892) were kindly provided by Dr. Michael Demetriou (UC Irvine, USA). *Rag1*^Cre^ transgenic mice, with a Cre recombinase gene introduced under the promoter of the Rag1 gene[34], were kindly provided by Dr. Marc Veldhoen (Instituto de Medicina Molecular, Portugal). Mice (males) with two floxed alleles for *Mgat1* or *Mgat2* were crossed with *Rag1*^Cre^ ones (females), with only one allele with the *Cre* gene, to generate *Rag1*^Cre^ with heterozygous floxed (both for *Mgat1* and *Mgat2*) progeny. These mice (females) were then crossed with homozygous floxed *Mgat1* or *Mgat2* ones (males) to generate *Rag1*^Cre^*Mgat1*^fl/fl^ (*Mgat1* cKO) and *Rag1*^Cre^*Mgat2*^fl/fl^ (*Mgat2* cKO) progeny. The littermates containing only the two floxed alleles (*Mgat1*WT and *Mgat2*WT) were used as controls. All mouse procedures were approved by the i3S ethics committee for animal experimentation under Portuguese regulation.

### Human thymocyte collection

#### Ethical Statement

Thymic specimens (173 to 578 days old) were obtained during routine thymectomy performed during pediatric corrective cardiac surgery at Hospital de Santa Cruz, Carnaxide, Portugal, after parent’s written informed consent, using thymic tissue that would be otherwise discarded. The study was approved by the Ethical Boards of Faculty of Medicine of the University of Lisbon and of Hospital de Santa Cruz, Carnaxide, Portugal.

### Preparation of human thymocyte populations

Total thymocytes were recovered through tissue dispersion and separation on a Ficoll-Paque PLUS (GE Healthcare) density gradient. Cells were then frozen in drop by drop added freezing medium (86% FCS, 14% DMSO), placed for 48H at -80° in isopropanol container and stored in liquid nitrogen until use, and cells were thawed immediately before use.

### Mice organ isolation

After CO2-mediated euthanasia, thymi and spleens were collected from age and sex matched mice and rinsed in PBS 2%FBS. Organs were macerated in a 70 μm nylon mesh, in order to generate single cell suspensions. For erythrocyte lysis, cells were incubated for 3 minutes at room temperature with 1x ACK (150 mM NH4Cl; 10 mM KHCO3; 0.1 nM Na2.EDTA), and washed in PBS 2%FBS. Cells were counted and stored in PBS 2%FBS on ice until further immediate use.

### Flow cytometry

Cell suspensions were stained with lectins and monoclonal antibodies from eBioscience and Biolegend, as described[7]. Briefly, for viability detection, cells were ressuspended in PBS and incubated with Fixable Viability Dye APC-Cy7 for 30 minutes on ice in the dark. For lectin staining, performed in murine and human thymocytes, 1x10^6^ cells were isolated and incubated with conjugated lectins for 30 minutes on ice, prior to antibody staining. Conjugated lectins (Vector labs) used were *Phaseolus Vulgaris* Leucoagglutinin (L-PHA), *Lycopersicon Esculentum* lectin (LEL), *Galanthus Nivalis* lectin (GNA), and *Sambucus Nigra* lectin (SNA). Antibodies were titrated and optimal concentrations were set, and cells were incubated for 30 minutes on ice in the dark. For intracellular staining, cells were fixed and permeabilized with the FoxP3 Fix/Perm buffer set. For TCRvß screen, cells were stained for specified markers, washed, and incubated for 30 minutes on ice in the dark with supplied antibodies suspensions. For galectin-1 binding, cells were stained for specified antibodies and incubated with recombinant galectin-1, as described[10]. Cells were analyzed in a FACS Canto II (BD Bioscience), and data was analyzed with the FlowJo v10 software.

### Neospora caninum infection

*N. caninum* tachyzoites (NcT) (Nc-1, ATCC® 50843) were propagated by serial passages in VERO cell cultures, maintained in Minimal Essential Medium (MEM) containing Earle’s salts (Sigma, St. Louis, MO, USA), supplemented with 10% fetal calf serum (BioWest, Nuaillé, France), L-Glutamine (2mM), Penicillin (100 IU/ml) and Streptomycin (100 µg/ml) (all from Sigma), in a humidified atmosphere with 5% CO2 at 37°C. Free parasites were obtained as previously described[26]. *N. caninum* challenge infections of 8-week old mice were performed by i.p. inoculation of 1*×*10^7^ freshly isolated NcT in 500 μL of PBS. Mouse weights and overall conditions were monitored daily until day 7 post infection. At euthanasia, spleens, lungs, and brains were collected for DNA isolation and parasite load quantification by qPCR, as described[26].

### Acute colitis induction

Colitis was induced in *Mgat1*WT, *Mgat1* cKO, *Mgat2*WT and *Mgat2* cKO 8-week old mice, using 2% DSS in water, for 7 days. Mouse weights, feces consistency, presence of blood in feces and rectum, and overall conditions were monitored daily. At euthanasia, colons and spleens were collected, for flow cytometry analysis. Colons were formalin-fixed and paraffin embedded and used for H&E staining, as described[7].

### Quantification and Statistical analysis

Statistical analyzes were performed using the GraphPad Prism 9 software. Further details of the statistics can be found in the Fig. legends. Multiple comparisons were done using the Kruskal-Wallis test. Significance values were computed using Mann-Whitney t-test. No statistical method was used to predetermine the sample size. All experiments were performed at least 2 times, with mice from independent progenies.

## Data Availability

All data analysed during this study are included in the manuscript, in the source data files. Further information and requests for resources and reagents should be directed and will be fulfilled to the Lead Contact Salome S Pinho.

## Acknowledgments

We would like to thank Dr. Michael Demetriou and Dr. Marc Veldhoen for providing *Mgat1*^fl/fl^ and *Mgat2*^fl/fl^, and *Rag1*^Cre^ mice, respectively.

---

**Supp. Fig. 1:**
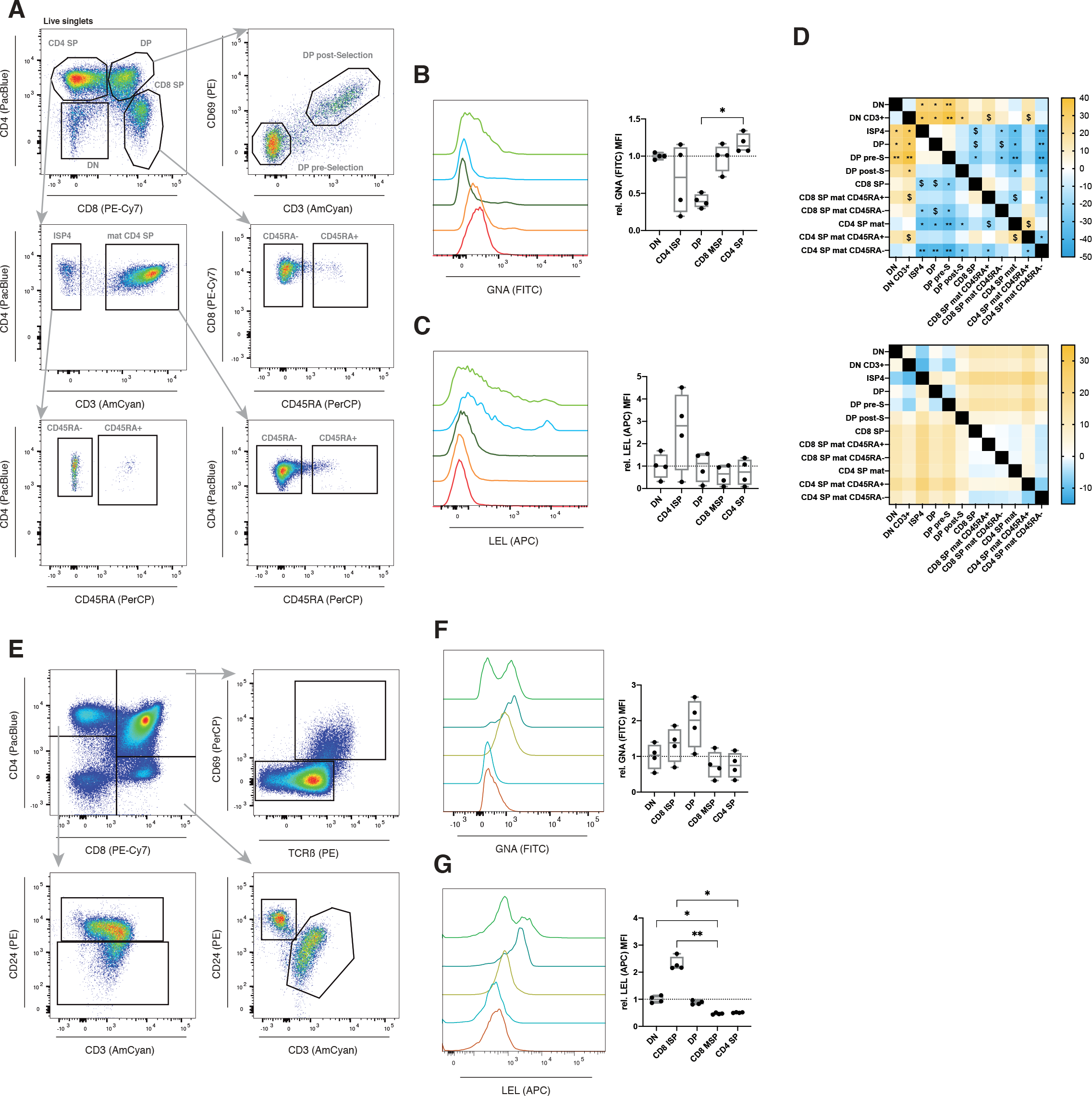
Glycoprofiling of human and murine thymocytes reveals developmentally regulated alterations. (A) and (F) Gating strategy for human and murine thymocyte subsets analysis, respectively. (B) and (G) GNA and (C) and (H) LEL binding levels for DN, CD4 ISP or ISP8, DP, CD8 MSP and CD4 MSP, for human (N = 4) and murine (N = 4) thymocytes, respectively. (E) and (I) Thymocyte population multiple comparison heatmap results for GNA (top) and LEL (bottom) binding levels, for human (N = 4) and mice (N = 4), respectively. Colorbar indicates the mean rank difference values. Kruskal-Wallis test, q-value $ < 0.1, * < 0.05, ** < 0.005 and *** < 0.001.

**Supp. Fig. 2:**
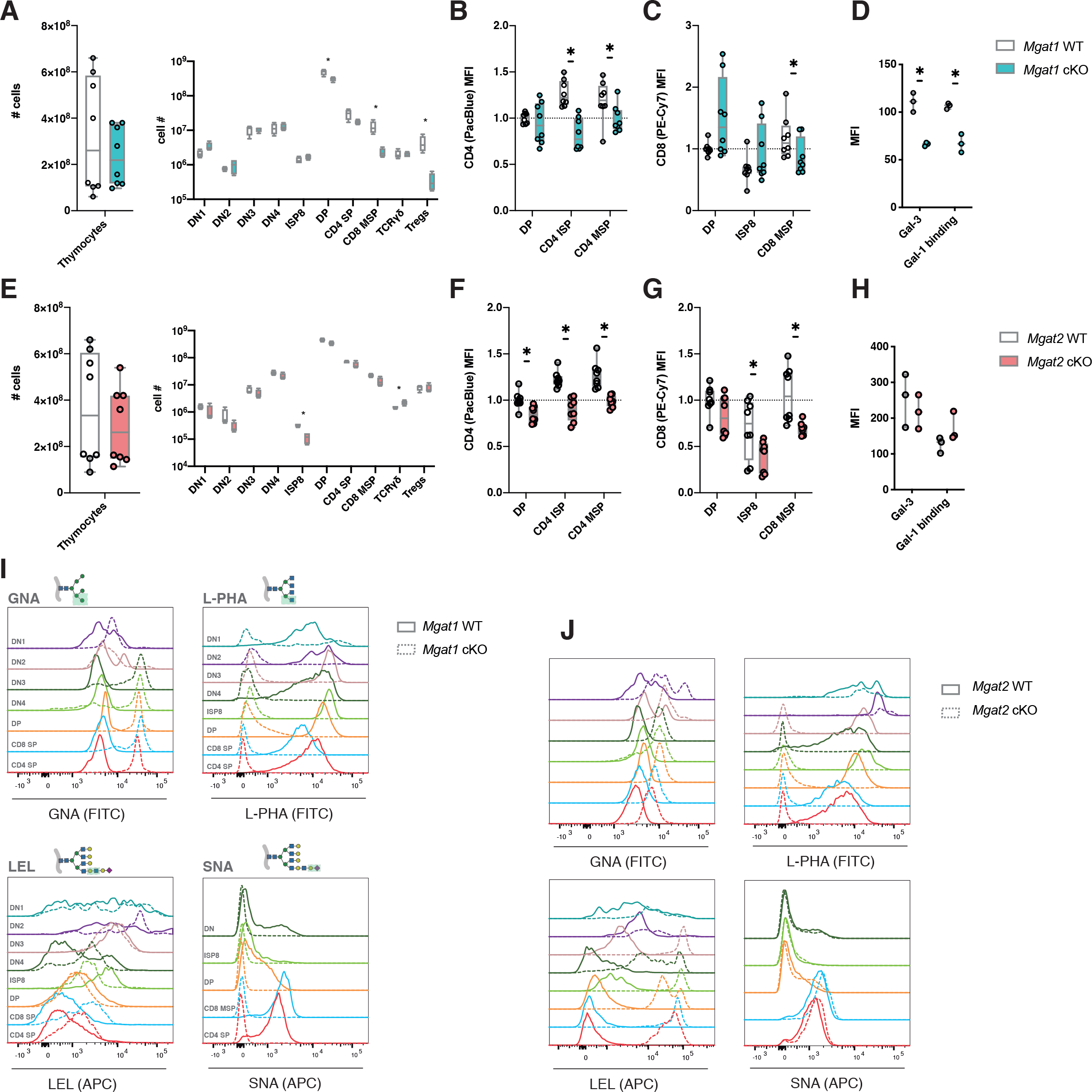
High-mannose restricted thymocytes fail to perform normal development. (A) and (E) Total thymocyte numbers in *Mgat1*WT (N = 8) and *Mgat1*cKO (N = 8), and *Mgat2*WT (N = 8) and *Mgat2*cKO (N = 8) mice, respectively. (B) and (F) CD4 and (C) and (G) CD8 coreceptor surface expression normalized levels (to the wildtype DP population) in *Mgat1*WT (N = 8) and *Mgat1*cKO (N = 8), and *Mgat2*WT (N = 8) and *Mgat2*cKO (N = 8) mice, respectively. (D) and (H) Endogenous Gal-3 and recombinant Gal-1 binding in DP thymocytes in *Mgat1*WT (N = 3) and *Mgat1*cKO (N = 3), and *Mgat2*WT (N = 3) and *Mgat2*cKO (N = 3) mice, respectively. (I) and (J) Representative histograms of lectin binding in in *Mgat1*WT (full lines) and *Mgat1*cKO (ticked lines), and *Mgat2*WT (full lines) and *Mgat2*cKO (ticked lines) mice, respectively.

**Supp. Fig. 3:**
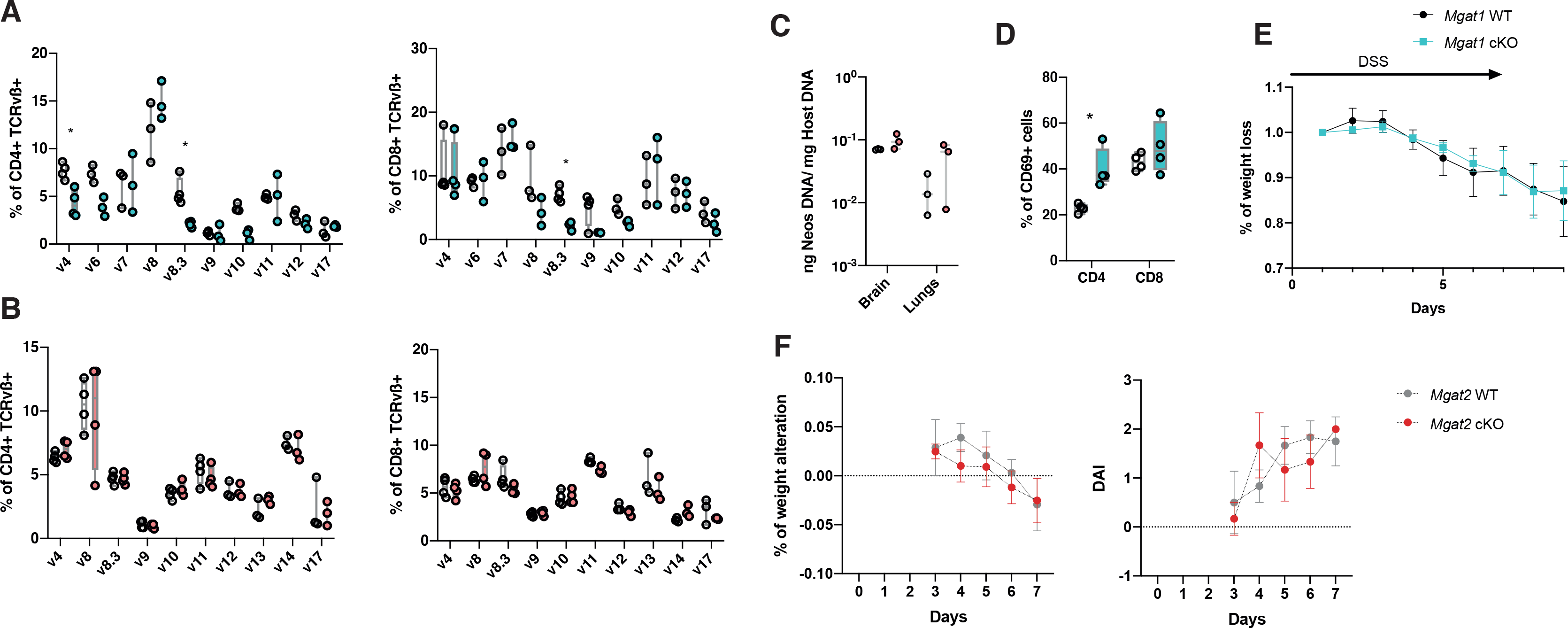
Absence of *Mgat1* in thymocytes impairs peripheral TCR repertoires and T cell responses. (A) and (B) Screen of TCRvß+ expressing cells, within splenic CD4 (left) and CD8 SP (right) in *Mgat1*WT (N = 4) and *Mgat1*cKO (N = 4), and *Mgat2*WT (N = 4) and *Mgat2*cKO (N = 4) mice, respectively. (C) *N. caninum* organ colonization determination, through the quantification of total parasite DNA in 1 mg of total host DNA, in *Mgat2*WT (N = 3) and *Mgat2*cKO (N = 3). (D) Quantification of the CD69 surface levels (MFI) for splenic CD4 and CD8 T cells, on the final day of infection. (E) Relative body weight loss of *Mgat1*WT (N = 4) and *Mgat1*cKO (N = 3) mice, upon DSS-induced colitis. (F) Relative body weight loss of *Mgat2*WT (N = 4) and *Mgat2*cKO (N = 4) mice, upon DSS-induced colitis (right) and disease Activity Index (DAI) scores for the DSS-induced colitis in *Mgat1*WT (N = 4) and *Mgat1*cKO (N = 3) (left).

**Supplementary Table 1.**

| REAGENT or RESOURCE | SOURCE | IDENTIFIER |
| --- | --- | --- |
| Antibodies |  |  |
| human CD4 (RPA-T4) eFlour 450 1:100 | eBioscience | 48-0049-41 |
| human CD8 (RPA-T8) PE-Cy7 1:400 | BD Pharmigen | 557750 |
| human CD69 (FN50) PE 1:100 | Biolegend | 310905 |
| human CD3 (OKT3) Brilliant Violet 510 1:100 | BD Pharmigen | 566779 |
| human CD45RA PerCP 1:100 | eBioscience | 45-0458-42 |
| mouse CD4 (RM4-5) eFluor 450 1:300 | eBioscience | 48-0042-80 |
| mouse CD8alpha (53-6.7) PE-Cy7 1:500 | eBioscience | 25-0081-81 |
| mouse CD69 (H1.2F3) PerCP-Cy5.5 1:200 | eBioscience | 45-0691-80 |
| mouse CD3 (17A2) Brilliant Violet 510 1:300 | Biolegend | 100233 |
| mouse CD24 (M1/69) PE 1:800 | eBioscience | 12-0242-81 |
| mouse TCRB (H57-597) PE 1:800 | eBioscience | 12-5961-81 |
| mouse CD25 (PC61.5) PE-Cy5 1:400 | eBioscience | 35-0251-80 |
| mouse CD44 (IM7) eFlour 506 1:100 | eBioscience | 69-0441-80 |
| mouse CD5 (53-7.3) PE-Cy5 1:200 | Biolegend | 100609 |
| mouse CD127 (A7R34 | eBioscience | 11-1271-81 |
| mouse TCRgd (eBioGL3) APC 1:400 | eBioscience | 17-5711-81 |
| mouse CD27 (LG.7F9) SuperBright 436 1:200 | eBioscience | 62-0271-80 |
| mouse FoxP3 (FJK-16S) APC 1:200 | eBioscience | 17-5773-80 |
| mouse CD62L (MEL-14) APC 1:200 | eBioscience | A14720 |
| mouse CD25 (PC61.5) PE 1:400 | eBioscience | 12-0251-81 |
| mouse CD19 (eBio1D3) APC 1:500 | eBioscience | 17-0193-80 |
| *Phaseolus vulgaris* Leucoagglutinin FITC<br>1:1000 | Vector<br>Laboratories | FL-1111-2 |
| *Galanthus nivalis* lectin FITC | Vector<br>Laboratories | FL-1241-2 |
| *Sambucus nigra* lectin Cy5 1:1000 | Vector<br>Laboratories | CL-1305-1 |
| *Lycopersicon esculentum* lectin DyLight 594<br>1:1000 | Vector<br>Laboratories | DL-1177-1 |

